# Comparative Burden of COVID-19 in Adults with Rheumatic Diseases and Immunocompetent Individuals: A Population-Based Cohort Study in Quebec, Canada, 2020-2023

**DOI:** 10.64898/2026.09.21.26363556

**Authors:** Sara Carazo, Inés Colmegna, Sasha Bernatsky, Paul R Fortin, Sonia Jean, Denis Talbot, Louis Rochette, Charles-Antoine Guay, Michael Libman, Michael Desjardins, Caroline Quach, Danuta M Skowronski, Gaston de Serres, Canadian Immunization Research Network

## Abstract

**Introduction:** Across major phases of the COVID-19 pandemic (March 2020-April 2023), we compared the burden of severe COVID-19 among individuals with systemic autoimmune rheumatic diseases (SARDs) versus immunocompetent individuals with and without comorbidities.

**Methods:** This population-based cohort study used administrative datasets to estimate crude and age-and sex-standardized SARS-CoV-2 and severe COVID-19 incidences among adults ≥18 years with SARDs, chronic comorbidities, or no comorbidities. Relative to the latter, we used Cox models to derive adjusted hazard ratios (aHRs) for infection, hospitalization, and intensive care unit admission or death (ICU/death) during the pre-vaccination (March–December 2020), vaccination pre-Omicron (December 2020– December 2021), and Omicron (December 2021–April 2023) periods.

**Results:** We included 81,434 adults with SARDs, and 2,020,356 with comorbidities versus 3,144,700 without comorbidities. Despite similar adjusted infection rates, the SARDS cohort had higher risks of hospitalization and ICU/death during the pre-vaccination period (aHRs: 3.4, 95%CI:2.8–4.1; and 4.0, 95%CI:2.8–5.7, respectively), persisting through the Omicron period (aHRs: 4.5, 95%CI:4.0–4.9; and 4.4, 95%CI:3.6–5.4, respectively). Among SARDS, rheumatoid arthritis, psoriatic arthritis and systemic lupus erythematosus contributed highest burden, with comparable hospitalization and mortality rates among younger rheumatoid arthritis versus healthy older adults. Highly immunosuppressive rituximab-or mycophenolate-treated patients had highest hospitalization and ICU/death risks, particularly during the Omicron period (aHRs: 11.9, 95%CI:9.9–14.3; and 17.6, 95%CI:13.0– 23.9, respectively).

**Conclusion:** Individuals with SARDs, especially receiving highly immunosuppressive therapies, experienced elevated COVID-19 burden and severe outcome risks throughout the pandemic, warranting their prioritized consideration during future public health emergencies.

## Introduction

During the COVID-19 pandemic, patients with immunosuppressive conditions or receiving immunosuppressive therapies were considered at increased risk of severe COVID-19 (1). Moreover, systemic autoimmune rheumatic diseases (SARDs) may be treated with a wide range of agents which vary substantially in their immunosuppressive potential (2). Disease-modifying antirheumatic drugs (DMARDs) increase susceptibility to viral infections through class-specific effects on antiviral immune pathways (3). These effects are dose-and context-dependent, often amplified by concomitant glucocorticoid use, and should be considered in conjunction with the underlying immune dysregulation of rheumatic diseases which also exacerbate the infection risk.

Patients with rheumatic diseases were generally considered as a single homogeneous high-risk group, in part due to the lack of robust empirical data quantifying the specific risk of severe COVID-19 by rheumatologic condition or individual immunomodulatory therapy. As a result, they were recommended to adopt more stringent preventive measures compared to the healthy population or even those with other chronic conditions (4). There remains a paucity of data on the burden of COVID-19 among patients with rheumatic diseases compared with other chronic conditions, or of the evolution in risk associated with treatment advancements, COVID-19 vaccine availability, and/or SARS-CoV-2 variant circulation.

Across major phases of the COVID-19 pandemic we quantified the burden of severe COVID-19 among a cohort of adults with SARDs relative to two immunocompetent cohorts including adults with other high-risk comorbidities and a reference cohort without comorbidities.

## Materials and Methods

### Study design

This cohort study in the province of Quebec, Canada, spanned March 2020 to April 2023 and was analyzed across three pandemic periods pre-defined in relation to major shifts in epidemiological patterns and/or public health interventions including: (A) March 1 to December 12, 2020 (epidemiological weeks 10-50); (B) December 27, 2020 to December 4, 2021 (epidemiological week 53 of 2020 to 48 of 2021); and (C) December 26, 2021 to April 27, 2023 (epidemiological week 52 of 2021 to 17 of 2023). Period A encompassed the first two pandemic waves due to ancestral SARS-CoV-2 marked by progressive expansion of testing capacity and implementation of public health measures including social distancing, restrictions on movement and gatherings, and mask use (5). Period B began with availability and prioritized rollout of COVID-19 vaccination, followed by expansion to the general population in March 2021. It was characterized by the emergence of SARS-CoV-2 variants of concern, primarily Alpha and Delta, universal access to nucleic acid amplification tests (NAAT) and adaptive public health measures based on the intensity of SARS-CoV-2 circulation. Period C was defined by emergence of the Omicron variant, which led to the largest and most intense wave of the pandemic, with extension of booster dose vaccination to the general population. Beginning in January 2022, NAAT access was restricted to individuals aged ≥65 years, healthcare workers, high-risk patients and those seeking care in acute-care facilities while rapid antigen tests became widely available in Quebec as elsewhere.

### Data sources

The cohorts were created using data from the Quebec Integrated Chronic Disease Surveillance System (QICDSS), extracted on March 31, 2023. In Quebec, all residents have access to universal publicly-funded healthcare, including medical consultations, tests or hospital stays. The QICDSS links multiple provincial health administrative databases, including the health insurance registry, hospitalization discharge database, physician fee-for-services billing claims database, the public prescription drug insurance database and the vital statistics registry (containing information on dates and causes of deaths) (6). Quebec’s public prescription drug insurance covers drugs for around 90% of individuals aged ≥65 years and approximately 37% of younger residents without private health care (including unemployed individuals, social assistance recipients, and many self-employed workers). Information on prescriptions reimbursed through private drug insurance plans, which are mandatory in Quebec for employed individuals and their families, was not available.

Using a unique personal identification number, participants were additionally linked to the provincial immunization registry and the laboratory database containing all NAAT for SARS-CoV-2.

### Study population

We included community-dwelling adults aged ≥18 years and listed in the provincial health insurance registry during the year preceding the study. Using data from the hospitalization discharge and fee-for-service physician billing claims databases between March 1996 and March 2020, we identified prevalent rheumatic and chronic conditions as of March 2020, at the start of the COVID-19 pandemic. The rheumatic disease cohort included individuals diagnosed with rheumatoid arthritis (RA), spondyloarthritis (SpA), psoriatic arthritis (PsA), systemic lupus erythematosus (SLE), systemic sclerosis (SSc), Sjögren’s syndrome (SS), inflammatory myositis (IM), vasculitis (VAS) or undifferentiated connective tissue disease (UCTD). Diseases were identified using validated algorithms when available, based on ICD-9 and ICD-10 codes recorded in hospitalization and in physician claims databases (Supplementary table 1). Analogous approaches were used to identify SARDs without validated algorithms. For individuals in the rheumatic disease cohort, data for 25 medications prescribed between March 2019 and March 2023 to treat SARDs were searched from the public drug insurance database using Drug Identification Number (DIN) codes (Supplementary table 2). The cohort with comorbidities included individuals with chronic conditions captured in the QICDSS that are not typically associated with immunodeficiency or the use of immunosuppressive therapies, including hypertension, diabetes mellitus (DM), chronic respiratory disease (CRD), cardiovascular or cerebrovascular disease (CVD), obesity and dementia. The cohort without comorbidities consisted of adults without any chronic condition identified by the QICDSS as being associated with increased risk of severe COVID-19 outcomes (Supplementary table 3) (7,8).

### Outcomes

SARS-CoV-2 infection was defined as a positive NAAT, reflecting both infection occurrence and access to testing. In Quebec, NAAT were universally accessible to symptomatic individuals and close contacts throughout 2020 and 2021. Only the first positive test was considered.

COVID-19 hospitalization was defined as a hospital admission lasting ≥24 hours with COVID-19 recorded as the primary reason for admission within 14 days following a positive SARS-CoV-2 test. Intensive care unit (ICU) admission was defined as any ICU stay occurring during a COVID-19 hospitalization. COVID-19 death was defined as a death for which COVID-19 was recorded as either the primary or a contributing cause of death in the provincial vital statistics registry, which does not contain SARS-CoV-2 testing information.

A composite outcome of ICU admission or COVID-19-related death (i.e., ICU/death) was used to capture very severe disease in multivariate analyses.

### Exposure

The cohorts were compared using cohort membership as the primary exposure. Within the rheumatic disease cohort and the cohort with comorbidities, the burden associated with each individual disease was described separately. Medications used in rheumatic diseases were grouped according to their presumed immunosuppressive potential. Three hierarchical treatment categories were defined: (drug-1) conventional or targeted immunomodulatory therapies, including methotrexate, leflunomide, and/or Janus kinase inhibitors (JAKi), with or without hydroxychloroquine, in the absence of therapies considered to confer higher levels of immunosuppression; (drug-2) glucocorticoid exposure, defined as prednisone use at any dose, with or without concomitant therapies; and (drug-3) highly immunosuppressive therapies, including rituximab, mycophenolate mofetil (MMF), belimumab, and/or cyclophosphamide, with or without other treatments.

### Statistical analyses

The prevalence of SARDs was calculated in the Quebec population at the start of the study period. We estimated crude and sex-and age-standardized incidence rates (IR) of SARS-CoV-2 infection, COVID-19 hospitalization and COVID-19 death per 100,000 person-years for each cohort, as well as according to specific rheumatic diseases, chronic diseases, and treatment categories across the three pandemic periods. We applied direct standardization using the age and sex distribution of the Quebec population in 2020 (9). Individuals contributed person-time from cohort entry until the earliest occurrence of a first positive SARS-CoV-2 NAAT, death, admission to a long-term care facility, or end of follow-up.

Cox proportional hazard models estimated adjusted hazard ratios (aHR) comparing the hazard of each outcome (infection, hospitalization, ICU/death) across exposure groups (cohorts, diseases or drug categories) with the reference cohort without comorbidities as the comparator. Separate models were fitted for each pandemic period and outcome. Calendar time was used as the underlying time scale. Models were adjusted for age, sex, number of comorbidities, geographic region, deprivation indices, eligibility for the public drug insurance plan and vaccination status, which was modeled as a time-varying covariate (from unvaccinated to five-dose vaccinated) during periods B and C. As recommended for explanatory models, competing events (deaths) were treated as censoring (10,11). Missing values for covariates, including comorbidity count and deprivation indices, were handled by including a distinct dummy category for missing values.

The mutually exclusive categories used for diseases, drug categories and covariates in multivariable analyses are described in Supplementary table 4.

The study cohorts included all Québec residents meeting the eligibility criteria, resulting in high statistical power (>80%) to detect absolute incidence differences of approximately 0.2% to 0.4% across the incidence range examined (3% to 15%).

### Ethics

This study was conducted under the legal mandate of the Quebec National Director of Public Health, under which the requirement for informed consent was waived. The study protocol was approved by the Research Ethics Board of the CHU de Québec-Université Laval.

## Results

### Population

At baseline, the rheumatic disease cohort comprised 81,434 individuals, the cohort with comorbidities 2,020,356 and the cohort without comorbidities 3,144,700 (Table 1). SARD patients were, as expected, predominantly women (64.9%), older than 50 years (76.4%), largely covered by the public drug insurance plan (70.4%) and commonly affected by comorbidities (75.0%). Individuals in the cohort with comorbidities had a similar age distribution, although women were less represented (51.2%), and the prevalence of other comorbidities was lower (57.4%). The cohort without comorbidities was substantially younger, with 66.2% aged 18-49 years.

**Table 1.** Characteristics of participants in each cohort.

|  | **Study cohorts** |  |  |
| --- | --- | --- | --- |
|  | Rheumatic disease cohort | Cohort with comorbidities | Cohort without comorbidities |
| Participants | 81,434 | 2,020,356 | 3,144,700 |
| Sex |  |  |  |
| Women | 52,869 (64.9) | 1,033,664 (51.2) | 1,455,188 (46.3) |
| Men | 28,565 (35.1) | 986,692 (48.8) | 1,689,512 (53.7) |
| Age, years |  |  |  |
| 18-49 | 19,236 (23.6) | 424,133 (21.0) | 2,083,336 (66.2) |
| 50-64 | 23,425 (28.8) | 612,154 (30.3) | 756,619 (24.1) |
| 65-79 | 27,127 (33.3) | 710,937 (35.2) | 278,457 (8.9) |
| ≥80 | 11,646 (14.3) | 273,132 (13.5) | 26,288 (0.8) |
| Public drug insurance | 57,336 (70.4) | 1,423,517 (70.5) | 1,377,859 (43.8) |
| Number of chronic diseases<sup>a</sup> |  |  |  |
| 0 | 0 (0.0) | 0 (0.0) | 3,144,700 (100.0) |
| 1 | 20,368 (25.0) | 861,197 (42.6) | 0 (0.0) |
| 2-4 | 40,326 (49.5) | 961,819 (47.6) | 0 (0.0) |
| 5-7 | 16,564 (20.3) | 175,240 (8.7) | 0 (0.0) |
| 8-10 | 3903 (4.8) | 21,333 (1.1) | 0 (0.0) |
| 11-16 | 273 (0.3) | 767 (0.0) | 0 (0.0) |
| Material deprivation index |  |  |  |
| Quintile 1 | 14,582 (17.9) | 327,184 (16.2) | 644,783 (20.5) |
| Quintile 2-4 | 44,092 (54.1) | 1,114,034 (55.1) | 1,782,756 (56.7) |
| Quintile 5 | 15,167 (18.6) | 407,445 (20.2) | 550,348 (17.5) |
| Unknown | 7593 (9.3) | 171,693 (8.5) | 166,813 (5.3) |
| Social deprivation index |  |  |  |
| Quintile 1 | 14,452 (17.7) | 357,194 (17.7) | 602,337 (19.2) |
| Quintile 2-4 | 44,747 (54.9) | 1,126,096 (55.7) | 1,796,820 (57.1) |
| Quintile 5 | 14,612 (18.0) | 365,373 (18.1) | 578,730 (18.4) |
| Unknown | 7593 (9.3) | 171,693 (8.5) | 166,813 (5.3) |

<sup>a</sup> Number of chronic diseases among the following 16: rheumatic diseases, high blood pressure, diabetes, cardiovascular diseases, chronic respiratory diseases, anemia, cancer, cerebrovascular disease, renal disease, liver disease, HIV, dementia, neurological disorder, mental disorders, substance abuse, obesity.

RA, SpA and SLE were the most common rheumatic diseases (prevalences of 432, 294 and 116/100,000 persons, respectively) (Table 2). Among individuals with RA covered by the public drug insurance plan, the most frequently prescribed therapies (alone or in combination with other DMARDs) were prednisone (10,978, 43.9%), methotrexate (10,751, 43.0%), hydroxychloroquine (8799, 35.2%) and TNF inhibitors (3380, 13.5%). Therapies categorized as highly immunosuppressive (drug-3), primarily MMF and rituximab, were most often prescribed for IM (19.4%), SSc (13.8%) and SLE (13.7%) (Table 2). Individuals with RA and SLE together accounted for 54.5% of all drug-3 prescriptions.

**Table 2.**
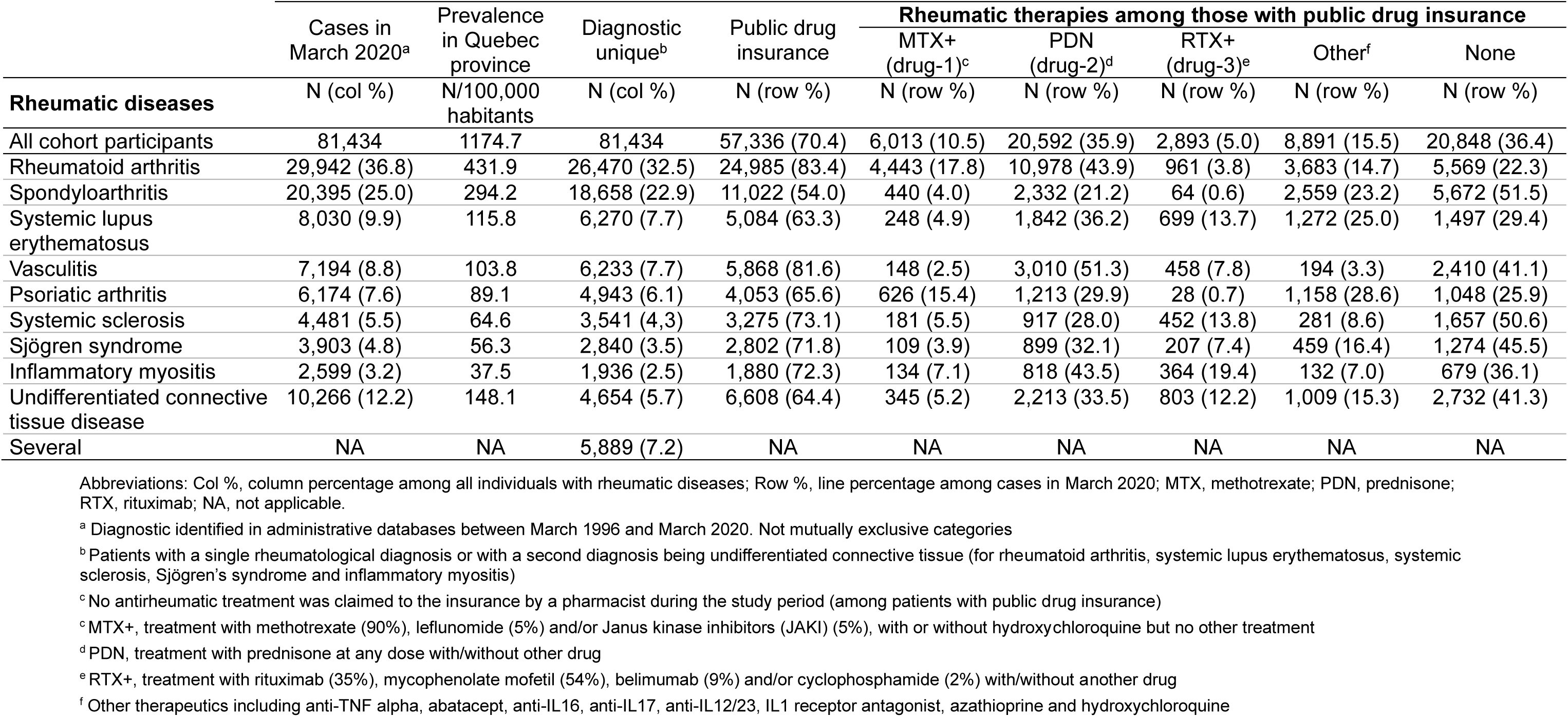
Rheumatic diseases and therapies.

### Incidence rates of COVID-19 outcomes

The epidemiological curves of the three cohorts closely mirrored the successive COVID-19 waves observed in Quebec, with the largest peak occurring in January 2022 following the emergence of the Omicron variant (Figure 1) (12). During the entire study period, 16.4% of individuals with SARDs had at least one documented SARS-CoV-2 infection, compared with 13.1% in the cohort with comorbidities and 12.1% in the cohort without comorbidities. Most infections occurred during the Omicron waves in period C, when 12.3%, 9.1% and 7.5%, respectively, of individuals of each cohort had a positive NAAT, compared to <3% in periods A and B. Among individuals with documented infection, hospitalization occurred in 19.0% (n=2,540) of individuals in the rheumatic disease cohort, compared with 12.6% (n=33,552) in the cohort with comorbidities and 1.2% (n=4,481) in the cohort without comorbidities.

**Figure 1.**
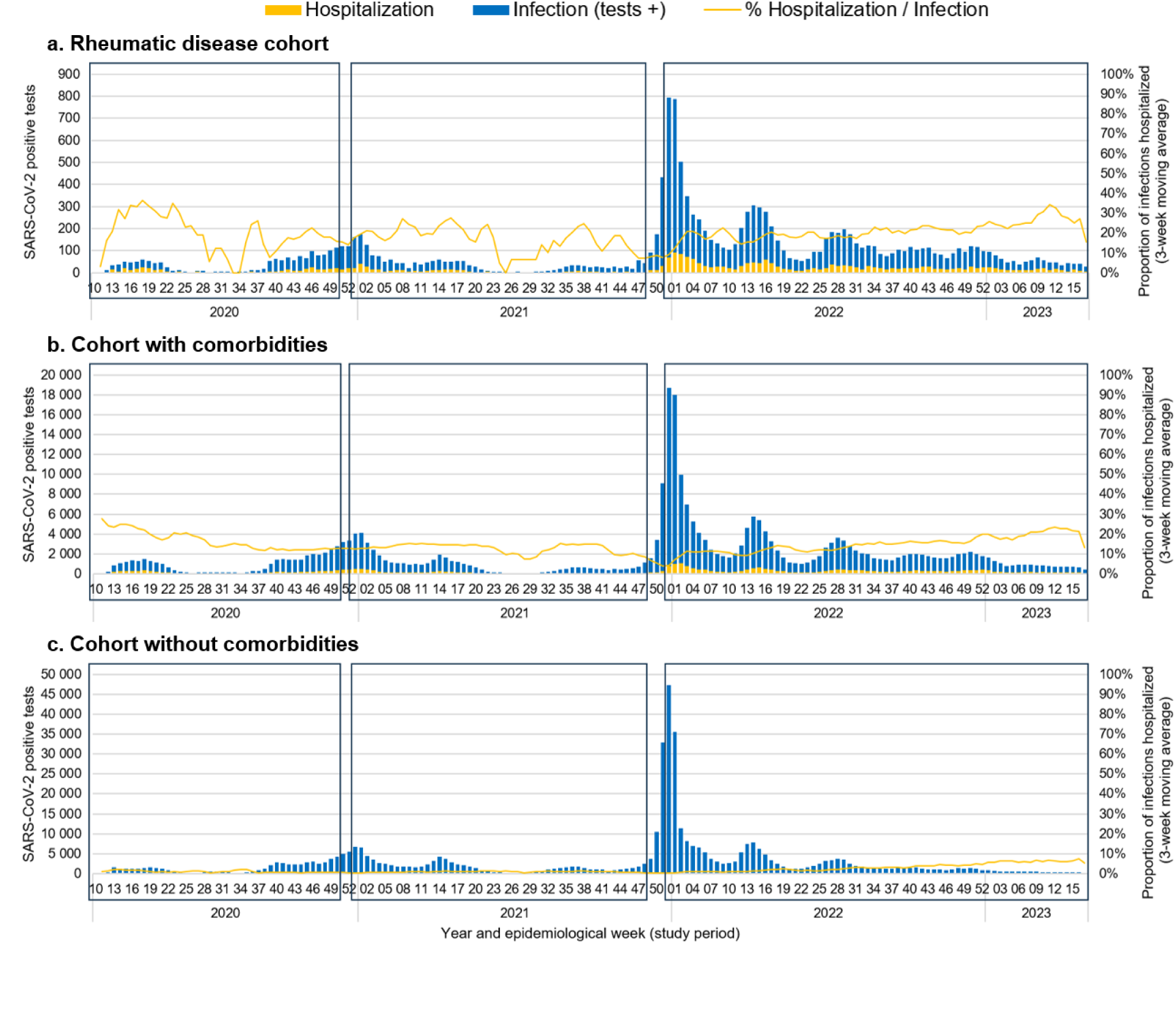
Temporal distribution of SARS-CoV-2 positive tests and hospitalizations by cohort and study period. Notes: The boxes in the figure represent the study periods; period A from week 2020-10 to week 2020-50, period B from week 2020-53 to 2021-48, and period C from week 2021-52 to 2023-17

**Figure 2.**
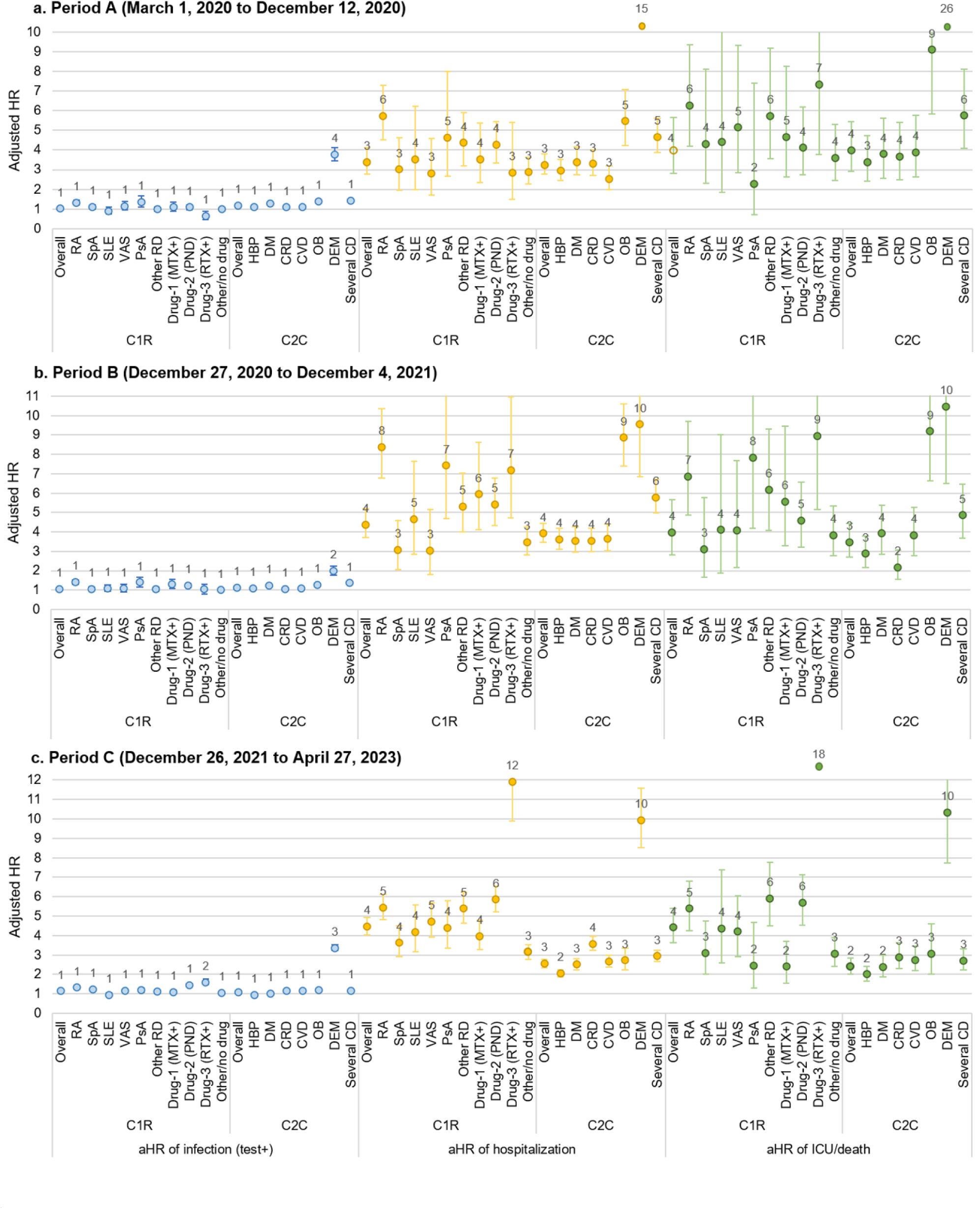
Adjusted hazard ratios of infection, hospitalization and death of individuals with rheumatic and chronic diseases, compared to healthy individuals. Abbreviations: C1R, rheumatic disease cohort; C2C, cohort with comorbidities; C3NC, cohort with no comorbidities; CI, confidence interval; CRD, chronic respiratory disease; CVD, cardiovascular diseases; DEM, dementia; DM, diabetes mellitus; HBP, high blood pressure; HR, hazard ratio; MTX+, treatment with methotrexate, leflunomide and/or Janus kinase inhibitors, with or without hydroxychloroquine but no other treatment; OB, obesity; PDN, treatment with prednisone at any dose with/without other drug; PsA, psoriatic arthritis; p-y, person-year; RA, rheumatoid arthritis; RTX+, treatment with rituximab, mycophenolate mofetil, belimumab and/or cyclophosphamide with/without another drug; SLE, systemic lupus erythematosus; SpA, spondyloarthritis; VAS, vasculitis. Note 1: Models included mutually exclusive categories compared cohort C1R and C2CD (model 1), rheumatic diseases and chronic diseases (model 2) and rheumatic treatments (model 3) to the cohort C3H (reference group for all models). They were adjusted for age, sex, region, number of comorbidities, vaccination status, public health insurance status and social and material deprivation indexes Note 2: Adjusted hazard ratios for individuals with dementia in the period A were: 15.0 (95%CI: 11.5 – 19.6) for hospitalization and 26.4 (95%CI: 17.6 – 39.6) for ICU/death (presented on the top of the panel a); and for individuals treated with MMF/rituximab was 17.6 (95%CI: 13.0 – 23.9) for ICU/death (presented on the top of the panel c)

Across all pandemic periods, standardized COVID-19 hospitalization and mortality rates were consistently higher in the rheumatic disease cohort than in the cohort with comorbidities and the cohort without comorbidities, despite broadly similar infection rates during periods A and B (Table 3). For example, hospitalization rates during period A were 319, 233 and 17 per 100,000 person-years, respectively, for each cohort, while the corresponding mortality rates were 101, 50 and 5. During the Omicron period C, infection, hospitalization, and mortality rates increased across all cohorts, with the rheumatic disease cohort continuing to experience the greatest burden of severe outcomes (ex. hospitalization rates of 1184, 443 and 83 per 100,000 person-years for each cohort) (Table 3).

**Table 3.**
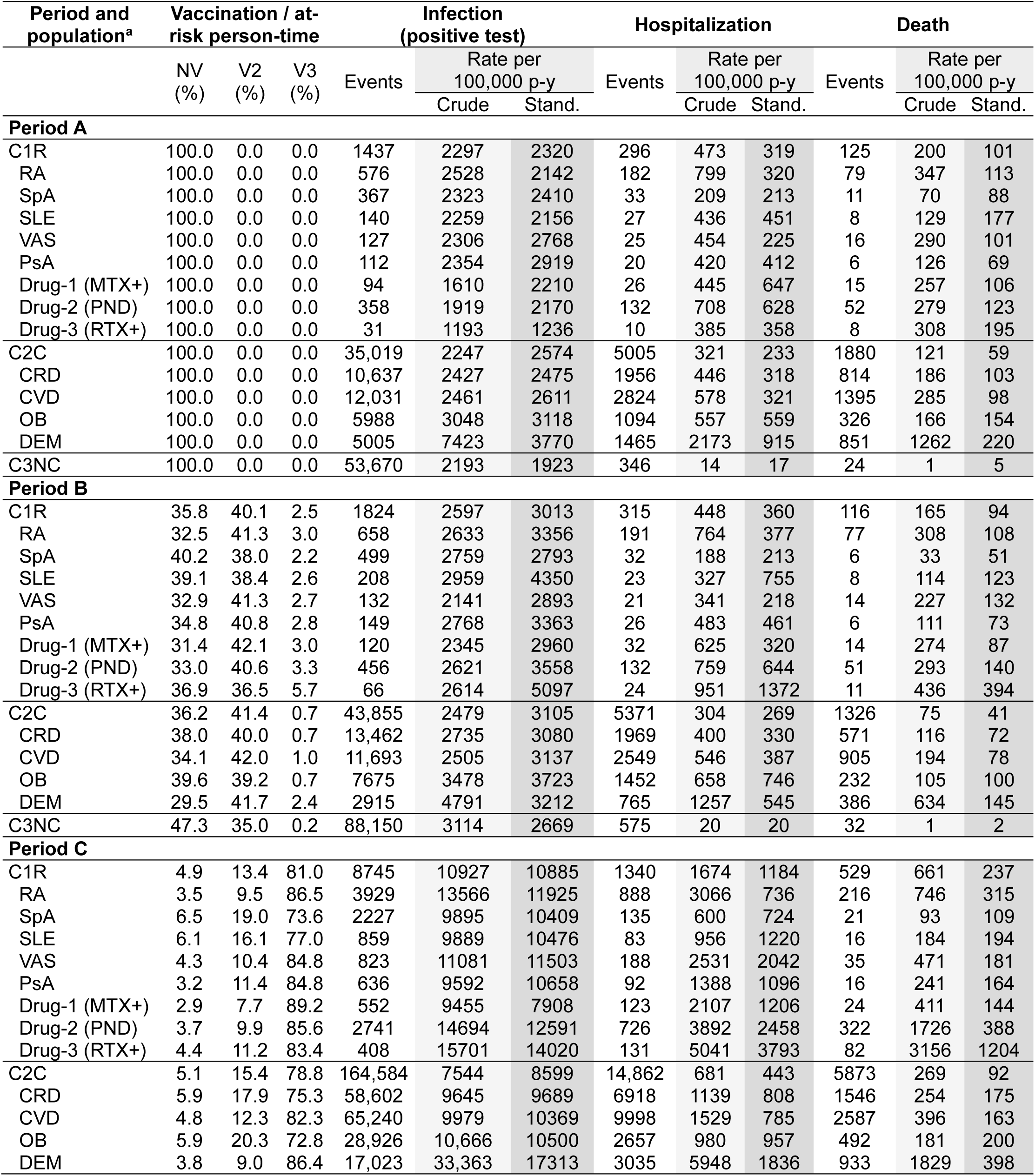
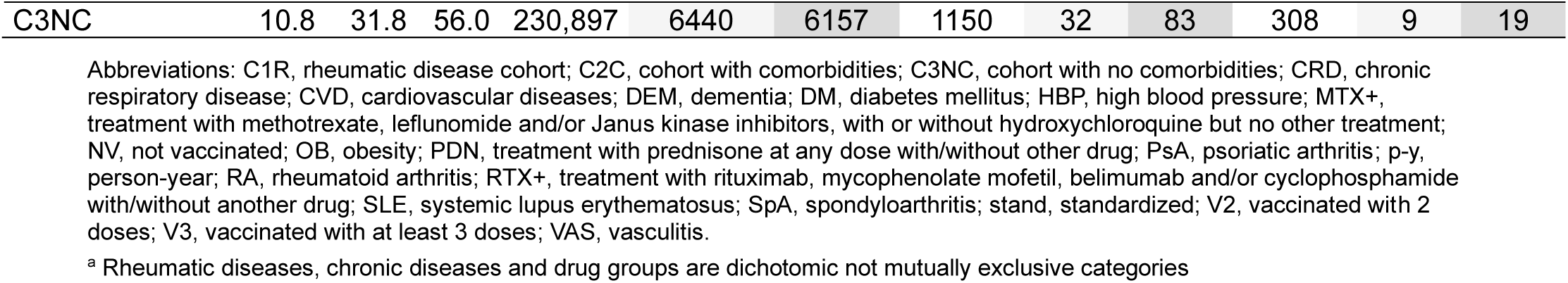
Crude and age-and sex-standardized rates of infection, hospitalization and death, by cohort and study period.

Considerable heterogeneity was observed across SARDs and treatment categories. Throughout the study period, patients with RA, SLE, and PsA generally experienced the highest hospitalization and mortality rates (Table 3). Age-stratified analyses showed that individuals with RA exhibited higher hospitalization and mortality rates than adults in the comparator cohorts across all age groups ≥50 years (Supplementary figures 1-3). During period C, the burden of severe COVID-19 among RA patients aged 50-64 years exceeded that observed among adults of the same age with CPD or CVD, as well as among adults aged 75-84 years without comorbidities (Supplementary figure 3).

Among drug categories, patients receiving highly immunosuppressive therapies, including rituximab/MMF (drug-3), consistently experienced the greatest burden of severe COVID-19. In period A, this group had infection rates nearly half those of the overall rheumatic disease cohort (1,236 versus 2320 per 100,000 person-years) but maintained elevated hospitalization and mortality rates (358 and 195 per 100,000 person-years, respectively). During period B, patients receiving prednisone (drug-2) or rituximab/MMF (drug-3) exhibited hospitalization and mortality rates nearly fourfold higher than those of the overall cohort. Disparities widened further in period C, when patients receiving rituximab/MMF experienced hospitalization and mortality rates of 3,793 and 1,204 per 100,000 person-years, respectively, compared with 1,206 and 144 among methotrexate-treated patients (drug-1) or 83 and 19 in the reference cohort without comorbidities (Table 3).

During the vaccination era (periods B and C), severe COVID-19 outcomes occurred disproportionately among unvaccinated individuals as shown by their higher incidences compared to the overall population (Supplementary Table 5; Table 3); for example, 360/100,000 person-years for SARDs overall versus 749 for those unvaccinated in period B and 1184 versus 2994, respectively, in period C. Although event counts were limited within some rheumatic disease subgroups, the relative between-and within-cohort patterns remained consistent with those observed in the overall population.

### Hazard Ratios of COVID-19 outcomes

Compared with the reference cohort without comorbidities, aHR for infection were generally similar across categories and periods (aHRs≈1) except for individuals with dementia (aHR=2.0 to 3.8) and for individuals receiving MMF or rituximab (drug-3) during period A (aHR=0.6; 95%CI:0.4–0.9) and period C (aHR=1.6; 95%CI:1.5–1.8) (Figure 3, Supplementary table 6). In contrast, and consistently relative to the reference cohort, individuals with SARDs exhibited comparable or higher aHR for hospitalization and ICU/death overall and by condition than those with chronic conditions except for obesity and dementia in periods A (aHRs≈3) and B (aHRs≈4). In period C, SARDs were associated with consistently higher aHR, overall (aHR=4.5; 95%CI:4.0-4.9 for hospitalization and aHR=4.4; 95%CI:3.6-5.4 for ICU/death versus aHR=2.5; 95%CI:2.3-2.8 and aHR=2.4; 95%CI:2.0-2.9 in the cohort with comorbidities) and by condition, except for dementia. Substantial heterogeneity was observed across rheumatic diseases, drug categories and periods. Among SARDs, RA was associated with the highest adjusted risks of hospitalization and ICU/death in all periods, except for ICU/death in period B, where the highest aHR was observed for PsA. However, differences between rheumatic diseases were less pronounced during period C. Among rheumatic therapies, MMF and rituximab (drug-3) were associated with the highest aHR for hospitalization and ICU/death, particularly in period C (11.9 [95%CI:9.9–14.3] and 17.6 [95%CI:13.0–23.9], respectively). By comparison, the corresponding aHR for the overall rheumatic disease cohort were 4.5 (95%CI:4.0–4.9) for hospitalization and 4.4 (95%CI:3.6– 5.4) for ICU/death.

## Discussion

In this large population-based cohort study conducted between 2020 and 2023, despite similar rates of laboratory-confirmed infections, patients with autoimmune rheumatic disease had higher rates of COVID-19 hospitalization and death than individuals with other high-risk chronic conditions or general population controls without comorbidities, with their excess risk having notably increased during the Omicron period. Across all periods, RA, the most prevalent rheumatologic diagnosis, accounted for the greatest burden of severe COVID-19 among SARDs. During post-vaccination periods B (pre-Omicron) and C (Omicron), individuals receiving highly immunosuppressive therapies (primary MMF or rituximab) or prednisone had substantially higher hospitalization and death rates than those treated with other immunomodulators or the broader SARD population.

During the first pandemic year, a Canadian study reported similar risk of infection in patients with immune-mediated inflammatory diseases (IMID) and the general population (13). We found similar risk of NAAT-confirmed SARS-CoV-2 infection between SARDs patients and non-immunocompromised populations during the first pandemic years, when NAAT testing was free and universal (12). This finding suggests comparable exposure and adherence to nonpharmaceutical interventions across cohorts despite public health recommendations for increased preventive measures in those with chronic conditions. Only patients receiving highly immunosuppressive therapies had reduced exposure during the first pandemic waves (aHR=0.6). Before Omicron emergence, the risk of hospitalization and death which are more robust outcomes than infection, have previously been reported 1.2 to 2.0 times higher in IMID than in matched general population cohorts, more importantly for RA, vasculitis and PsA (12–14). In our study, SARD patients experienced hospitalization and ICU admission/death rates 1.3 to 2.3 times higher than patients with comorbidities, and 3 to 4 times higher than healthy individuals. RA and PsA patients were generally associated with the highest aHR.

Our findings indicate that the greater risk of severe outcomes persisted during the Omicron period despite vaccine coverage exceeding 95% and similar aHR of infection between cohorts. A large Greek cohort study of RA patients in 2022 reported hospitalization and death rates of 32 and 6 per 1000 person-years, respectively, with incidence rate ratios of 2.0 and 1.8 compared with matched general population controls (14). We observed comparable IR of 31 and 7 per 1000 person-years among RA patients; however, aHR were 2.4 and 2.5 relative to the chronic disease cohort and exceeded 7 when compared to the healthy cohort without comorbidities.

DMARDs may increase susceptibility to viral infections through class-specific but convergent effects on key antiviral immune pathways (3). Conventional synthetic DMARDs such as methotrexate can impair proliferation and function of antigen-presenting cells and lymphocytes, thereby attenuating both innate sensing and adaptive responses to viral pathogens (15). Targeted synthetic DMARDs, particularly JAKi, can disrupt type I and III interferon signaling pathways that are central to early antiviral defense, potentially impairing viral containment and delaying viral clearance (16). Biologic DMARDs further modulate host defenses by inhibiting pathways critical for host protection; for example, B cell–depleting therapies such as rituximab markedly impair humoral immunity and memory B-cell formation, leading to reduced neutralizing antibody responses and prolonged viral shedding (17). Although we were unable to reliably assess each DMARD category separately, our observations support differential risks of severe COVID-19 across treatment classes. The greater risk in patients using highly immunosuppressive therapies or prednisone has been previously observed. During the pre-Omicron period, several studies reported higher risk of hospitalization and death among patients treated with B-cell depletion (rituximab) than among those under conventional synthetic (cs) DMARDs (15–18). Among csDMARDs, methotrexate, hydroxychloroquine, and TNF inhibitors carried the lowest risk and prednisolone the highest (18). During the Omicron period, rituximab was also the DMARD associated with the highest risk of COVID-19 hospitalization and death (odds ratios (OR) of 6.1 and 12.1, respectively) in a cohort of RA patients (14) and in a cohort of SARD patients (hospitalization OR of 2.2) (19). We observed hospitalization and mortality aHR of 12 and 18 for patients under MMF or rituximab compared with the healthy-reference cohort. The high rates of severe COVID-19 associated with prednisone use in our study are in line with observations of increased COVID-19 hospitalization risk during the Omicron period (19) and previous findings of association of steroid therapy and serious infection (20,21).

This study has potential limitations. Algorithms used to identify rheumatic and other chronic diseases, although not formally validated in Quebec administrative databases, have demonstrated high specificity in other jurisdictions (22,23). We lacked information on certain high-risk conditions for COVID-19 complications, such as tuberculosis, smoking status or some forms of immunocompromise (7,24), potentially leading to misclassification within the cohort without comorbidities and underestimation of the relative burden of the rheumatic disease cohort and the cohort with comorbidities. Medication data were limited to prescriptions covered under the public drug insurance plan, and no DMARDs were identified in 36 % of covered patients with rheumatic diseases, limiting our ability to conduct drug-specific analyses. Furthermore, although DMARDs were prescribed during the study period and prior to SARS-CoV-2 infection, some treatments, particularly prednisone, may have been used intermittently, discontinued or administered at low doses, potentially attenuating their observed impact on COVID-19 severity. Finally, hospitalization practices evolved throughout the pandemic period and may have varied by comorbidity status. However, such differences in clinical practice would not have impact on COVID-19 mortality rates, which were consistent with the observed patterns in hospitalization.

Our study highlights the high absolute and relative rates of severe COVID-19 in SARDs, across three pandemic years representing epidemiological and clinical changes including advances in COVID-19 therapy, vaccine introduction, changing testing practices and variant evolution. This excess risk was amplified during the Omicron period, despite targeted measures such as the use of Nirmatrelvir/Ritonavir since April 2022 for early outpatient treatment in high-risk patients (25) or the enhancement of a third vaccine dose recommended in immunosuppressed populations (26).

While prior studies often compared rheumatic disease patients with matched general population controls, we performed direct comparisons with two population-based cohorts, including individuals with high-risk chronic conditions prioritized for COVID-19 prevention and treatment, and with stratifications by disease and age-group providing insights relevant to public health and emergency preparedness and response. In December 2020, immunization programs in Quebec and other jurisdictions such as the US and UK, proposed vaccine prioritization in the context of insufficient supply based on risk of infection (healthcare workers) and age, considered the main risk factor for hospitalization and death (25–27). Adults <65 years with chronic conditions were only considered after universal vaccination of all individuals>60 years, and without specific mention of immunosuppressed individuals. The greater risk of severe outcomes for younger patients with SARDs and other chronic diseases warrants their prioritized consideration during subsequent epidemics or pandemics.

In conclusion, individuals with SARDs experienced a persistently elevated burden of COVID-19 hospitalization, ICU admission and death from 2020 to 2023, exceeding that observed in non-immunocompromised individuals, including older adults and those with other high-risk conditions such as respiratory or cardiovascular diseases. These finding support prioritizing SARD patients, and particularly those who are immunocompromised, alongside other of the highest risk patients during public health emergencies.

## Data Availability

Data are not publicly available due to privacy restrictions. Data are held by the Quebec Ministry of Health and the Régie de l'assurance maladie du Québec (RAMQ), and access is restricted to authorized personnel. Requests for data access should be directed to the Ministère de la Santé et des Services sociaux du Québec. The study code is available from the authors upon reasonable request.

## Acknowledgments

We thank Katia Giguère (Institut national de santé publique du Québec) for her contribution to the statistical analyses.

## Data sharing

Data are not publicly available due to privacy restrictions. Data are held by the Québec Ministry of Health and the Régie de l’assurance maladie du Québec (RAMQ), and access is restricted to authorized personnel. Requests for data access should be directed to the Ministère de la Santé et des Services sociaux du Québec. The study code is available from the authors upon reasonable request.

## References

1. Antinori A, Bausch-Jurken M. The burden of COVID-19 in the immunocompromised patient: Implications for vaccination and needs for the future. J Infect Dis 2023;228:S4-12.

2. Furer V, Rondaan C, Heijstek MW, et al. 2019 update of EULAR recommendations for vaccination in adult patients with autoimmune inflammatory rheumatic diseases. Ann Rheum Dis 2020;79:39–52.

3. Winthrop KL, Mariette X. To immunosuppress: whom, when and how? That is the question with COVID-19. Ann Rheum Dis 2020;79:1129–31.

4. Hazlewood GS, Pardo JP, Barnabe C, Schieir O, Barber CEH, Bernatsky S, et al. Canadian Rheumatology Association recommendation for the use of COVID-19 vaccination for patients with autoimmune rheumatic diseases. J Rheumatol 2021;48:1330–9.

5. Institut national de santé publique du Québec. COVID-19 (coronavirus). 2023. Ligne du temps COVID-19 au Québec. Available at: https://www.inspq.qc.ca/covid-19/donnees/variants

6. Blais C, Jean S, Sirois C, et al. Quebec Integrated Chronic Disease Surveillance System (QICDSS), an innovative approach. Chronic Dis Inj Can 2014;34:226–35.

7. Centers for Disease Control and Prevention. Covid. 2025. People with certain medical conditions and COVID-19 risk factors. Available at: https://www.cdc.gov/covid/risk-factors/index.html

8. Simard M, de Montigny C, Sonia J, Fortin E. Impact of comorbidities on the risk of death and hospitalization among confirmed cases of COVID-19 during the first months of the pandemic in Québec. Institut national de santé publique du Québec; 2020. Available at: https://www.inspq.qc.ca/sites/default/files/publications/3082-impact-comorbidities-risk-death-covid-19.pdf

9. Institut de la Statistique du Québec. Population and demography. 2025. Population of Quebec by age and gender, 1971–2025. Available at: https://statistique.quebec.ca/en/produit/tableau/697#tri_pop=30

10. Austin PC, Lee DS, Fine JP. Introduction to the Analysis of Survival Data in the Presence of Competing Risks. Circulation 2016;133:601–9.

11. Austin PC, Fine JP. Practical recommendations for reporting Fine-G ray model analyses for competing risk data. Stat Med 2017;36:4391–400.

12. Institut national de santé publique du Québec. INSPQ. 2026. Données Covid-19 au Québec. Available at: https://www.inspq.qc.ca/covid-19/donnees

13. Eder L, Croxford R, Drucker AM, et al. Understanding COVID-19 risk in patients with immune-mediated inflamatory diseases: a population based analysis of SARS-CoV-2 testing. Arthritis Care Res 2023;75:317–25.

14. Bournia VK, Fragoulis GE, Mitrou P, et al. Outcomes of COVID-19 Omicron variant in patients with rheumatoid arthritis: a nationwide Greek cohort study. Rheumatology 2024;63:1130–8.

15. Cronstein BN, Aune TM. Methotrexate and its mechanisms of action in inflammatory arthritis. Nat Rev Rheumatol 2020;16:145–54.

16. Zhang Q, Bastard P, Liu Z, et al. Inborn errors of type I IFN immunity in patients with life-threatening COVID-19. Science 2020;370:eabd4570.

17. Avouac J, Drumez E, Hachulla E, et al. COVID-19 outcomes in patients with inflammatory rheumatic and musculoskeletal diseases treated with rituximab: a cohort study. Lancet Rheumatol 2021;3:e419–26.

18. McKeigue P, Porter D, Hollick R, et al. Risk of severe COVID-19 in patients with inflammatory rheumatic diseases treated with immunosuppressive therapy in Scotland. Scand J Rheumatol 2023;52:412–7.

19. Patel NJ, Srivatsan S, Kowalski EN, et al. Patients with systemic autoimmune rheumatic diseases remain at risk for hospitalisation for COVID-19 infection in the Omicron era (2022– 2024): a retrospective cohort study. RMD Open 2025;11:e005114.

20. Doran MF, Crowson CS, Pond GR, O’Fallon WM, Gabriel SE. Predictors of infection in rheumatoid arthritis. Arthritis Rheum 2002;46:2294–300.

21. De Almeida ALB, Guimarães MFBR, Da Costa Pinto MR, al. Predictors of serious infections in rheumatoid arthritis—a prospective Brazilian cohort. Adv Rheumatol 2024;64:23.

22. Widdifield J, Bernatsky S, Paterson JM, et al. Accuracy of Canadian health administrative databases in identifying patients with rheumatoid arthritis: A validation study using the medical records of rheumatologists. Arthritis Care Res 2013;65:1582–91.

23. Eder L, Widdifield J, Rosen CF, et al. Identifying and characterizing psoriasis and psoriatic arthritis patients in Ontario administrative data: A population-based study from 1991 to 2015. J Rheumatol. 1 nov 2020;47(11):1644–51. doi:10.3899/jrheum.190659

24. Adas MA, Bechman K, Russell MD, et al. Risk of infection in patients with early inflammatory arthritis: results from a large UK prospective observational cohort study. Rheumatology 2025;64:5647–55.

25. INESSS. COVID-19: Antivirals, anticoagulants and immunomodulators. 2022. Available at: https://www.inesss.qc.ca/fileadmin/doc/INESSS/COVID-19/2024-GUO_COVID-19_EN.pdf

26. Comité sur l’immunization du Québec. Avis portant sur la pertinence d’une dose additionnelle de vaccin contre la COVDI-19 pour les personnes ayant une immunodépression. 2021. Available at: https://www.inspq.qc.ca/publications/3163-pertinence-dose-additionnelle-vaccin-covid-19-immunodeprimes

