## Supplemental material for "Comparative Burden of COVID-19 in Adults with Rheumatic Diseases and Immunocompetent Individuals: A Population-Based Cohort Study in Quebec, Canada, 2020-2023"

### Supplementary material

Supplementary table 4. Codification of the variables included in the multivariate models………..7

Supplementary table 5. Age- and sex-standardized incidence rates of infection, hospitalization and death among unvaccinated individuals, by cohort and study period………………………...…8

Supplementary table 6. Crude and adjusted hazard ratios of COVID-19 infection, hospitalization and death by cohort, disease, treatment class and period………………………….…………..10

Supplementary figure 1. Infection, hospitalization and death unadjusted rates by cohort, disease and age group during the period A…………………………………………………………….……..12

Supplementary figure 2. Infection, hospitalization and death unadjusted rates by cohort, disease and age group during the period B…………………………………………………..……………13

Supplementary figure 3. Infection, hospitalization and death unadjusted rates by cohort, disease and age group during the period C……………………………………………………….………14

**Supplementary table 1. Algorithms and diagnostic codes to identify the cohorts of individuals with rheumatic and chronic diseases**

| Diagnostic | ICD-9 and ICD-10 codes | Case definition |
|---|---|---|
| **1. Diseases to define rheumatic disease cohort** | | |
| Rheumatoid arthritis | 714.x<br>M05.x, M06.x | One hospitalization since 1996 or three physician billing claims, with at least one claim submitted by a rheumatologist, orthopedist or internal medicine specialist within two years |
| Spondyloarthritis | 720.x<br>M45.x | One hospitalization since 1996 or three physician billing claims, with at least one claim submitted by a rheumatologist or internal medicine specialist within two years |
| Psoriatic arthritis | 606.0<br>L40.5, M07.0, M07.1, M07.2, M07.3, M09.0 | One hospitalization since 1996 or three physician billing claims submitted by any physician |
| Systemic lupus erythematosus | 710.0<br>M32.1, M32.8, M32.9 | One hospitalization since 1996 or three physician billing claims, with at least one claim submitted by a rheumatologist or internal medicine specialist within two years |
| Systemic sclerosis | 710.1<br>M34.x | One hospitalization since 1996 or three physician billing claims, with at least one claim submitted by a rheumatologist or internal medicine specialist within two years |
| Sjögren's syndrome | 710.2<br>M35.0 | One hospitalization since 1996 or three physician billing claims, with at least one claim submitted by a rheumatologist, orthopedist or internal medicine specialist within two years |
| Inflammatory myositis | 710.3, 710.4<br>M33.x | One hospitalization since 1996 or three physician billing claims, with at least one claim submitted by a rheumatologist or internal medicine specialist within two years |
| Vasculitis | 446.x<br>M30.0, M30.1, M31.3, M31.4, M31.5, M31.7 | One hospitalization since 1996 or three physician billing claims, with at least one claim submitted by a rheumatologist or internal medicine specialist within two years |
| Undifferentiated connective tissue disease | 710.8, 710.9<br>M35.9 | One hospitalization since 1996 or three physician billing claims, with at least one claim submitted by a rheumatologist or |

| | | |
|---|---|---|
| | | internal medicine specialist within two years |
| **2. Chronic conditions to define cohort with comorbidities** | | |
| High blood pressure | 401.x, 402.x–405.x, 437.2<br>I10.x, I11.x–I13.x, I15.x, I67.4 | One hospitalization since 1996 or two physician billing claims within 730 days |
| Diabetes mellitus | 250.0-250.9<br>E10.0-E10.9, E11.0-E11.9, E13.0-E13.9, E14.0-E14.9 | One hospitalization since 1996 or two physician billing claims within 730 days (among individuals 20 years or older) |
| Chronic respiratory diseases | 490x–505.x, 506.4, 508.1, 508.8[a]<br>I27.8[c], I27.9[c], J40.x-J47.x, J60.x-J64.x,<br>J65.x, J66.x, J67.x,<br>J68.4, J70.1, J70.3 | One hospitalization since 1996 or three physician billing claims within 1096 days (chronic pulmonary obstructive disease)<br>One hospitalization or two physician billing claims within 730 days (asthma) |
| Cardiovascular diseases<br>Valvular disease<br>Cardiac arrhythmias<br>Myocardial infarction<br>Congestive heart failure<br>Cerebrovascular disease | 394.x–397.x, 424.x, 746.3–746.6, V42.2, V43.3<br>426.0, 426.7, 426.9, 427.0–427.4, 427.6–427.9, 785.0, V45.0, V53.3<br>410.x, 412.x<br>402.1, 404.1, 428.x<br>I05.x–I08.x, I09.1, I09.8, I34.x–I39.x, Q23.0–Q23.3, Q23.8, Q23.9, Z95.2, Z95.3, Z95.4<br>I44.1–I44.3, I45.6, I45.9, I47.x–I49.x, R00.0, R00.1, R00.8, T82.1, Z45.0, Z95.0<br>I21.x, I22.x, I25.2<br>I09.9, I11.0, I13.0, I13.2, I25.5, I42.0, I42.5–I42.9, I43.x, I50.x, P29.0<br>362.34, 430.x-438.x<br>G45.x, G46.x, I60.x-I69.x | One hospitalization since 1996 or two physician billing claims within 365 days |
| Obesity | 278.0<br>E66.x | One hospitalization since 2013 (10-year lookback) or two physician billing claims within 730 days and separated more than 30 days |
| Dementia | 290.x, 294.1, 331.0, 331.2<br>F00.x-F03.x, F05.1, G30.x, G31.1 | One hospitalization since 1996 or three physician billing claims within 730 days and separated more than 30 days (40 years or older) |

### Supplementary table 2. Therapeutics for systemic rheumatic diseases

| Drug type | Drug | Drug identification number (DIN) codes |
|---|---|---|
| Conventional synthetic DMARDs | Methotrexate | 1915; 321397; 410195; 614327; 614335; 614343; 632619; 1907204; 1911457; 2099705; 2170655; 2170671; 2170698; 2182750; 2182777; 2182947; 2182955; 2182963; 2182971; 2244798; 2320029; 2320037; 2320045; 2327236; 2398427; 2417626; 2419173; 2422166; 2422174; 2422182; 2422190; 2422204; 2454750; 2454769; 2454777; 2454831; 2454858; 2454866; 2454874; 2464365; 2491311; 2491338; 2491346; 2491354; 2491362; 2509067 |
| | Leflunomide | 2241888; 2241889; 2256495; 2256509; 2261251; 2261278; 2283964; 2283972; 2288265; 2288273; 2309327; 2309335; 2319225; 2319233; 2351668; 2351676; 2415828; 2415836 |
| | Azathioprine | 4596; 337854; 2231491; 2236799; 2236819; 2242148; 2242907; 2243371; 2343002; 99100908 |
| | Mycophenolate mofetil | 2192748; 2237484; 2242145; 2313855; 2320630; 2348675; 2352559; 2352567; 2364883; 2370549; 2371154; 2378574; 2379996; 2380382; 2383780; 2386399; 2389754; 2432625; 2433680; 2457369; 2457377; 2522233 |
| | Hydroxychloroquine | 1928287; 2017709; 2246691; 2252600; 2311011; 2424991; 2491427; 2511886; 2519348 |
| Anti-CD20 monoclonal antibodies | Rituximab | 2241927; 2457350; 2498316; 99114051; 2513447; 2495724; 2478382; 2478390 |
| Anti-BAFF monoclonal antibody | Belimumab | 2370050; 2370069; 2470470; 2470489 |
| Anti-TNF-α agents | Adalimumab | 2258595; 2452999; 2459302; 2459310; 2473097; 2473100; 2474263; 2492156; 2492164; 2502380; 2502399; 2502402; 2502674; 2502682; 2505258; 2511045; 2511053; 2511061; 2523760; 2523779; 2523949; 2523957; 2523965; 99100385; 99113852; 99113853; 99113854; 99113855; 99113856; 99113885; 99113886; 99113887; 99113962; 99113963; 99113964; 99113965; 99113967; 99113968; 99113969; 99113971; 99113972; 99113973; 99114033; 99114034; 99114035; 99114039; 99114040; 99114041; 99114042; 99114043; 99114046; 99114047; 99114048; 99114049; 99114050 |

| | | |
|---|---|---|
| | Etanercept | 2242903; 2274728; 2455323; 2455331; 2462850; 2462869; 2462877; 99100373 |
| | Infliximab | 2244016; 2419475; 2470373; 2496933; 2511584; 99101167; 99108493; 99108693; 99108694; 99108893; 99109093 |
| | Golimumab | 2324776; 2324784; 2413175; 2413183; 2417472 |
| | Certolizumab pegol | 2331675; 2465574 |
| Anti-IL6 receptor antibodies | Tocilizumab | 2350092; 2350106; 2350114; 2424770; 2483327; 99114013; 99114014; 99114015 |
| | Sarilumab | 2460521; 2460548; 2472961; 2472988 |
| Anti-IL17 pathway inhibitors | Secukinumab | 2438070; 99101215 |
| | Ixekizumab | 2455102; 2455110 |
| | Brodalumab | 2473623; 99107893 |
| Anti-IL12/23 monoclonal antibody | Ustekinumab | 2320673; 2320681; 2459671 |
| IL1 receptor antagonist | Anakinra | 2245913 |
| T-cell costimulation modulator | Abatacept | 2282097; 2402475 |
| Janus kinase inhibitors (JAKI) | Tofacitinib | 2423898; 2470608; 2480786 |
| | Baricitinib | 2480018 |
| | Upadacitinib | 2495155; 2520893 |
| Alkylating agents | Cyclophosphamide | 13544; 13552; 13749; 262676; 344877; 344885; 344915; 373753; 602833; 602949; 99101374; 2241795; 2241796; 2241797; 2241798; 2241799 |
| Systemic glucocorticoids | Prednisone | 21695; 156876; 210188; 232378; 252417; 271373; 312770; 550957; 598194; 607517; 610623; 99100987; 99100988 |

Abbreviations: BAFF, B-cell activating factor; DIN, drug identification number; DMARDs, disease-modifying antirheumatic drug; IL, interleukin; JAKI, Janus kinase inhibitors; TNF, tumor necrosis factor

### Supplementary table 3. List of chronic diseases the absence of which defined the cohort without comorbidities

| Comorbidities associated with an increased risk of severe COVID-19[1] | Diseases included from the QICDSS |
|---|---|
| High blood pressure | High blood pressure |
| Diabetes | Diabetes, uncomplicated<br>Diabetes, complicated |
| Chronic heart conditions | Myocardial infarction<br>Congestive heart failure<br>Valvular disease<br>Cardiac arrhythmias<br>Peripheral vascular disorders |
| Cerebrovascular disease | Cerebrovascular disease |
| Chronic kidney disease | Chronic kidney disease |
| Chronic liver disease | Chronic liver disease |
| Chronic respiratory diseases | Chronic pulmonary disease<br>Pulmonary circulation disorder |
| Cancer | Cancer without metastasis<br>Metastatic cancer |
| Dementia and other neurological conditions | Dementia<br>Neurological disorder<br>Paralysis |
| Fluid and electrolyte disorder | Fluid and electrolyte disorder |
| Hypothyroidism | Hypothyroidism |
| Hemoglobin blood disorders | Anemia |
| HIV infection | HIV |
| Immunocompromised condition | Rheumatic diseases |
| Mental health conditions | Psychoses<br>Depression |
| Obesity | Obesity |
| Substance use disorders | Alcohol abuse<br>Drug abuse |
| Coagulopathy | Coagulopathy |
| Weight loss | Weight loss |
| Ulcer disease | Ulcer disease |

Abbreviations: QICDSS, Quebec integrated chronic disease surveillance system

[1] Impact of chronic comorbidities on hospitalization, intensive care unit admission and death among adult vaccinated and unvaccinated COVID-19 confirmed cases during the Omicron wave - Marc Simard, Véronique Boiteau, Élise Fortin, Sonia Jean, Louis Rochette, Pierre-Luc Trépanier, Rodica Gilca, 2023

### Supplementary table 4. Codification of the variables included in the multivariate models

| Cox model | Exposure or covariables | Categories |
|---|---|---|
| 1 | Cohort (exposure) | C1R = rheumatic disease cohort<br>C2C = cohort with comorbidities<br>C3NC = cohort without comorbidities |
| 2 | Disease (exposure) | RA = rheumatoid arthritis[a]<br>SpA = spondyloarthritis<br>SLE = systemic lupus erythematosus[a]<br>VASC = vasculitis<br>PsA = psoriatic arthritis<br>OtherRheu = other rheumatic disease or several rheumatic diseases<br>HBP = high blood pression<br>DM = diabetes mellitus<br>CRD = chronic respiratory diseases<br>CVD = cardiovascular diseases<br>OB = obesity<br>DEM = dementia<br>SeveralCD = several chronic diseases among HBP, DM, CRD, CVD, OB and DEM |
| **3** | Rheumatic drug categories (exposure) | Drug-1 = methotrexate, leflunomide and/or Janus kinase inhibitors (JAKI) (with or without hydroxychloroquine but no other treatment)<br>Drug-2 = prednisone (with or without another drug but excluding drugs of group 3)<br>Drug-3 = rituximab, mycophenolate mofetil, belimumab and/or cyclophosphamide (with or without another drug) |
| 1 to 3 | Age in March 2020 | 18-49, 50-64, 65-79, ≥80 years |
| 1 to 3 | Sex | Male, female |
| 1 to 3 | Number of comorbidities | 0-1, 2-3, 4-5, ≥6 comorbidities |
| 1 to 3 | Socio-sanitary region | Regions 03, 04, 05, 06, 12, 13, 14, 15, 16 and one category for those with <5% of population (regions 01, 02, 07, 08, 09, 10, 11, 17, 18); 10 categories |
| 1 to 3 | Material deprivation index | Quintile 1, 5, 2-4 or unknown; 3 categories |
| 1 to 3 | Social deprivation index | Quintile 1, 5, 2-4 or unknown; 3 categories |
| 1 to 3 | Public drug insurance status | Oui (all or part of the period), non |
| 1 to 3 (periods B and C) | Vaccination (time-varying variable) | 0 to 5 doses; 6 categories |

[a] Including those having also the diagnosis of undifferentiated connective tissue disease

**Supplementary table 5. Age- and sex-standardized incidence rates of infection, hospitalization and death among unvaccinated individuals, by cohort and study period**

| Period and population[a] | Infection (positive test) | | Hospitalization | | Death | |
|---|---|---|---|---|---|---|
| | Events | Rate per 100,000 p-y | Events | Rate per 100,000 p-y | Events | Rate per 100,000 p-y |
| **Period A** | | | | | | |
| C1R | 1437 | 2320 | 296 | 319 | 125 | 101 |
| RA | 576 | 2142 | 182 | 320 | 79 | 113 |
| SpA | 367 | 2410 | 33 | 213 | 11 | 88 |
| SLE | 140 | 2156 | 27 | 451 | 8 | 177 |
| VAS | 127 | 2768 | 25 | 225 | 16 | 101 |
| PsA | 112 | 2919 | 20 | 412 | 6 | 69 |
| Drug-1 (MTX+) | 94 | 2210 | 26 | 647 | 15 | 106 |
| Drug-2 (PND) | 358 | 2170 | 132 | 628 | 52 | 123 |
| Drug-3 (RTX+) | 31 | 1236 | 10 | 358 | 8 | 195 |
| C2C | 35,019 | 2574 | 5005 | 233 | 1880 | 59 |
| CRD | 10,637 | 2475 | 1956 | 318 | 814 | 103 |
| CVD | 12,031 | 2611 | 2824 | 321 | 1395 | 98 |
| OB | 5988 | 3118 | 1094 | 559 | 326 | 154 |
| DEM | 5005 | 3770 | 1465 | 915 | 851 | 220 |
| C3NC | 53,670 | 1923 | 346 | 17 | 24 | 5 |
| **Period B** | | | | | | |
| C1R | 1249 | 5290 | 220 | 749 | 80 | 224 |
| RA | 414 | 5566 | 129 | 799 | 52 | 257 |
| SpA | 365 | 4991 | 31 | 597 | <5 | * |
| SLE | 157 | 7482 | 17 | 1185 | 6 | 259 |
| VAS | 82 | 4235 | 11 | 385 | 8 | 215 |
| PsA | 95 | 6041 | 16 | 856 | <5 | * |
| Drug-1 (MTX+) | 79 | 5712 | 20 | 674 | 10 | 245 |
| Drug-2 (PND) | 295 | 6063 | 90 | 1320 | 29 | 273 |
| Drug-3 (RTX+) | 44 | 7616 | 13 | 1604 | 7 | 539 |
| C2C | 32,115 | 5553 | 4202 | 583 | 987 | 104 |
| CRD | 9554 | 5458 | 1386 | 710 | 379 | 179 |
| CVD | 9017 | 5583 | 1950 | 824 | 678 | 188 |
| OB | 5602 | 6747 | 1133 | 1583 | 148 | 227 |
| DEM | 1944 | 6225 | 506 | 1242 | 272 | 349 |
| C3NC | 69,855 | 4403 | 532 | 40 | 28 | 4 |
| **Period C** | | | | | | |
| C1R | 442 | 10560 | 118 | 2994 | 29 | 704 |
| RA | 176 | 14521 | 73 | 220 | 20 | 1102 |
| SpA | 104 | 8896 | 15 | 2366 | <5 | * |
| SLE | 49 | 8454 | 9 | 2040 | <5 | * |
| VAS | 48 | 16123 | 19 | 6393 | <5 | * |
| PsA | 26 | 14268 | 5 | 2921 | <5 | * |
| Drug-1 (MTX+) | 27 | 16474 | 13 | 5227 | 5 | 2771 |
| Drug-2 (PND) | 142 | 18247 | 54 | 6740 | 8 | 666 |
| Drug-3 (RTX+) | 19 | 19078 | 5 | 3270 | <5 | * |
| C2C | 8918 | 7546 | 1933 | 1431 | 521 | 350 |
| CRD | 3102 | 9354 | 715 | 2440 | 185 | 677 |
| CVD | 3400 | 10014 | 1000 | 2205 | 310 | 576 |
| OB | 1541 | 10930 | 342 | 3362 | 59 | 758 |
| DEM | 734 | 21768 | 279 | 5483 | 117 | 1462 |
| C3NC | 14,232 | 3570 | 302 | 172 | 47 | 48 |

Abbreviations: C1R, rheumatic disease cohort; C2C, cohort with comorbidities; C3NC, cohort without comorbidities; CRD, chronic respiratory disease; CVD, cardiovascular diseases; DEM, dementia; DM, diabetes mellitus; HBP, high blood pressure; MTX+, treatment with methotrexate, leflunomide and/or Janus kinase inhibitors, with or without hydroxychloroquine but no other treatment; OB, obesity; PDN, treatment with prednisone at any dose with/without other drug; PsA, psoriatic arthritis; p-y, person-years; RA, rheumatoid arthritis; RTX+, treatment with rituximab, mycophenolate mofetil, belimumab and/or cyclophosphamide with/without another drug; SLE, systemic lupus erythematosus; SpA, spondyloarthritis; VAS, vasculitis

* Number of events lower than 5

[a] Rheumatic diseases, chronic diseases and drug groups are dichotomic (yes/non), not mutually exclusive categories

### Supplementary table 6. Crude and adjusted hazard ratios of COVID-19 infection, hospitalization and death by cohort, disease, treatment class and period

| Period and population[a] | Infection (positive test) | | Hospitalization | | Death | |
|---|---|---|---|---|---|---|
| | Crude HR (95% CI) | Adjusted HR (95%CI) | Crude HR (95% CI) | Adjusted HR (95%CI) | Crude HR (95% CI) | Adjusted HR (95%CI) |
| **Period A** | | | | | | |
| C1R | 1.0 (0.9–1.0) | 1.0 (1.0–1.1) | 33.5 (28.7–39.1) | 3.4 (2.8–4.1) | 86.6 (65.9–113.8) | 4.0 (2.8–5.7) |
| RA | 1.2 (1.1–1.3) | 1.3 (1.2–1.5) | 57.7 (47.9–69.5) | 5.7 (4.5–7.3) | 146.3 (107.5–199.1) | 6.3 (4.2–9.4) |
| SpA | 1.0 (0.9–1.2) | 1.1 (1.0–1.2) | 11.7 (7.8–17.8) | 3.0 (2.0–4.6) | 29.7 (16.5–53.6) | 4.3 (2.3–8.1) |
| SLE | 1.0 (0.8–1.2) | 0.9 (0.7–1.1) | 19.0 (10.9–33.0) | 3.5 (2.0–6.2) | 41.0 (17.8–94.1) | 4.4 (1.8–10.5) |
| VAS | 1.0 (0.8–1.2) | 1.1 (0.9–1.4) | 26.6 (16.6–42.8) | 2.8 (1.7–4.6) | 110.7 (64.5–190.0) | 5.1 (2.8–9.3) |
| PsA | 1.1 (0.9–1.3) | 1.3 (1.1–1.7) | 26.0 (15.2–44.3) | 4.6 (2.7–8.0) | 26.0 (8.2–82.5) | 2.3 (0.7–7.4) |
| Other/several RD | 0.9 (0.8–1.0 | 1.0 (0.9–1.1) | 30.7 (23.5–40.2) | 4.4 (3.2–5.9) | 79.7 (53.3–119.2) | 5.7 (3.6–9.2) |
| Drug-1 (MTX+) | 0.9 (0.8–1.1) | 1.1 (0.9–1.3) | 40.1 (26.9–59.7) | 3.5 (2.3–5.4) | 122.4 (72.3–207.4) | 4.7 (2.6–8.3) |
| Drug-2 (PND) | 1.1 (1.0–1.2) | 1.1 (1.0–1.2) | 61.5 (50.1–75.5) | 4.3 (3.4–5.4) | 136.7 (97.1–192.4) | 4.1 (2.7–6.2) |
| Drug-3 (RTX+) | 0.6 (0.4–0.9) | 0.6 (0.4–0.9) | 31.7 (16.9–59.4) | 2.8 (1.5–5.4) | 163.0 (86.5–306.9) | 7.3 (3.8–14.4) |
| Other/no drug | 1.1 (1.0–1.1) | 1.0 (0.9–1.1) | 23.2 (19.0–28.3) | 2.9 (2.3–3.6) | 59.9 (43.4–82.5) | 3.6 (2.4–5.3) |
| C2C | 1.0 (1.0–1.0) | 1.2 (1.1-1.2) | 22.7 (20.4–25.3) | 3.2 (2.8–3.8) | 58.8 (46.7–73.9) | 4.0 (2.9–5.4) |
| HBP | 0.8 (0.7–0.8) | 1.1 (1.1–1.1) | 7.3 (6.3–8.4) | 2.9 (2.5–3.5) | 13.0 (9.9–17.2) | 3.4 (2.4–4.7) |
| DM | 0.9 (0.9–1.0) | 1.3 (1.2–1.3) | 7.5 (6.2–9.1) | 3.4 (2.7–4.2) | 11.8 (8.3–16.8) | 3.8 (2.6–5.6) |
| CRD | 1.0 (1.0–1.0) | 1.1 (1.0–1.1) | 6.8 (5.7–8.1) | 3.3 (2.7–4.0) | 9.9 (7.0–13.9) | 3.7 (2.5–5.4) |
| CVD | 0.9 (0.8–0.9) | 1.1 (1.0–1.1) | 7.5 (6.1–9.2) | 2.5 (2.0–3.2) | 18.8 (13.5–26.2) | 3.9 (2.6–5.8) |
| OB | 1.5 (1.4–1.6) | 1.4 (1.3–1.4) | 9.9 (7.8–12.5) | 5.5 (4.2–7.1) | 18.4 (12.3–27.6) | 9.1 (5.8–14.2) |
| DEM | 2.5 (2.3–2.7) | 3.8 (3.4–4.1) | 66.0 (52.3–83.2) | 15.0 (11.5–19.6) | 229.0 (164.6–318.8) | 26.4 (17.6–39.6) |
| Several CD | 1.2 (1.2–1.2) | 1.4 (1.4–1.5) | 45.2 (40.5–50.5) | 4.7 (3.9–5.6) | 124.4 (98.7–156.8) | 5.7 (4.1–8.1) |
| C3NC | Reference | Reference | Reference | Reference | Reference | Reference |
| **Period B** | | | | | | |
| C1R | 0.8 (0.8–0.8) | 1.1 (1.1–1.1) | 14.9 (1.7–16.2) | 3.9 (3.4–4.4) | 26.8 (22.7–31.7) | 3.5 (2.7–4.4) |
| RA | 0.8 (0.8–0.9) | 1.4 (1.3–1.5) | 37.8 (31.9–44.9) | 8.4 (6.8–10.4) | 80.6 (62.3–104.4) | 6.9 (4.9–9.7) |
| SpA | 0.9 (0.8–1.0) | 1.0 (1.0–1.1) | 7.7 (5.2–11.4) | 3.1 (2.0–4.6) | 13.8 (7.7–24.9) | 3.1 (1.7–5.8) |
| SLE | 1.0 (0.8–1.1) | 1.1 (0.9–1.3) | 15.1 (9.3–24.5) | 4.7 (2.8–7.6) | 24.2 (11.3–51.6) | 4.1 (1.9–9.0) |
| VAS | 0.7 (0.6–0.8) | 1.1 (0.9–1.3) | 13.7 (8.2–22.9) | 3.0 (1.8–5.2) | 42.5 (23.6–76.6) | 4.1 (2.2–7.7) |
| PsA | 0.9 (0.7–1.1) | 1.4 (1.2–1.7) | 22.7 (14.5–35.4) | 7.4 (4.7–11.7) | 52.7 (29.3–94.9) | 7.8 (4.2–14.6) |
| Other/several RD | 0.8 (0.7–0.8) | 1.0 (0.9–1.2) | 19.8 (15.3–25.5) | 5.3 (4.0–7.0) | 47.6 (33.7–67.3) | 6.2 (4.1–9.3) |
| Drug-1 (MTX+) | 0.8 (0.6–0.9) | 1.3 (1.1–1.5) | 30.6 (21.4–43.7) | 5.9 (4.1–8.6) | 66.8 (40.9–108.9) | 5.6 (3.3–9.4) |
| Drug-2 (PND) | 0.8 (0.8–0.9) | 1.2 (1.1–1.3) | 36.3 (29.7–44.2) | 5.4 (4.3–6.8) | 72.1 (53.4–97.4) | 4.6 (3.2–6.6) |
| Drug-3 (RTX+) | 0.8 (0.7–1.1) | 1.0 (0.8–1.3) | 46.6 (31.0–70.1) | 7.2 (4.7–11.0) | 120.4 (71.9–201.7) | 8.9 (5.1–15.5) |
| Other/no drug | 0.8 (0.8–0.9) | 1.0 (0.9–1.1) | 14.9 (12.4–17.9) | 3.5 (2.8–4.3) | 34.2 (26.1–44.6) | 3.8 (2.8–5.3) |
| C2C | 0.8 (0.8–0.9) | 1.0 (1.0–1.1) | 22.0 (19.2–25.2) | 4.4 (3.7–5.2) | 48.2 (38.8–60.0) | 4.4 (3.3–5.9) |
| HBP | 0.6 (0.6–0.6) | 1.1 (1.0–1.1) | 5.6 (5.0–6.3) | 3.6 (3.1–4.2) | 7.1 (5.7–8.9) | 2.9 (2.2–3.8) |
| DM | 0.8 (0.8–0.8) | 1.2 (1.2–1.3) | 5.7 (5.0–6.3) | 3.5 (3.0–4.2) | 9.0 (6.8–11.7) | 3.9 (2.9–5.4) |
| CRD | 0.9 (0.9–0.9) | 1.1 (1.0–1.1) | 5.8 (5.0–6.7) | 3.5 (3.0–4.2) | 5.2 (3.9–7.0) | 2.2 (1.6–3.1) |
| CVD | 0.8 (0.7–0.8) | 1.1 (1.0–1.1) | 7.5 (6.4–8.8) | 3.7 (3.0–4.4) | 13.7 (10.6–17.7) | 3.8 (2.8–5.3) |
| OB | 1.3 (1.2–1.3) | 1.3 (1.2–1.3) | 13.9 (11.9–16.3) | 8.9 (7.4–10.6) | 20.2 (15.3–26.7) | 9.2 (6.6–12.7) |
| DEM | 0.9 (0.8–1.0) | 2.0 (1.8–2.2) | 20.5 (15.0–28.1) | 9.5 (6.9–13.3) | 45.5 (29.6–70.0) | 10.5 (6.5–16.9) |
| Several CD | 0.8 (0.8–0.8) | 1.4 (1.3–1.4) | 28.1 (25.7–30.6) | 5.8 (5.0–6.7) | 55.0 (46.5–65.0) | 4.9 (3.7–6.4) |
| C3NC | Reference | Reference | Reference | Reference | Reference | Reference |
| **Period C** | | | | | | |
| C1R | 1.6 (1.6–1.7) | 1.1 (1.1–1.2) | 52.0 (48.1–56.1) | 4.5 (4.0–4.9) | 69.7 (60.0–81.0) | 4.4 (3.6–5.4) |
| RA | 2.0 (1.9–2.0) | 1.3 (1.3–1.4) | 91.7 (83.7–100.4) | 5.4 (4.8–6.1) | 127.3 (107.2–151.2) | 5.4 (4.3–6.8) |
| SpA | 1.5 (1.4–1.5) | 1.2 (1.2–1.3) | 14.8 (12.0–18.2) | 3.6 (2.9–4.5) | 16.1 (10.7–24.3) | 3.1 (2.0–4.8) |
| SLE | 1.5 (1.4–1.6) | 1.0 (0.9–1.0) | 23.8 (18.1–31.4) | 4.2 (3.2–5.6) | 31.1 (18.8–51.4) | 4.4 (2.6–7.4) |

| | | | | | | |
|---|---|---|---|---|---|---|
| VAS | 1.6 (1.5–1.7) | 1.1 (1.1–1.2) | 68.5 (57.6–81.4) | 4.7 (3.9–5.7) | 85.4 (61.7–118.2) | 4.2 (2.9–6.0) |
| PsA | 1.3 (1.2–1.4) | 1.2 (1.1–1.3) | 32.9 (25.3–42.9) | 4.4 (3.3–5.8) | 24.5 (13.0–46.0) | 2.5 (1.3–4.7) |
| Other/several RD | 1.6 (1.5–1.6) | 1.1 (1.1–1.2) | 48.5 (42.9–54.9) | 5.4 (4.7–6.2) | 70.5 (56.4–88.2) | 5.9 (4.5–7.8) |
| Drug-1 (MTX+) | 1.4 (1.3–1.5) | 1.1 (1.0–1.2) | 63.4 (52.9–76.1) | 4.0 (3.3–4.8) | 52.8 (35.1–79.6) | 2.4 (1.6–3.7) |
| Drug-2 (PND) | 2.1 (2.0–2.2) | 1.4 (1.4–1.5) | 112.1 (102.0–123.3) | 5.9 (5.2–6.6) | 149.5 (124.8–179.1) | 5.7 (4.5–7.1) |
| Drug-3 (RTX+) | 2.2 (2.0–2.5) | 1.6 (1.5–1.8) | 148.5 (124.9–176.6) | 11.9 (9.9–14.3) | 274.5 (208.4–361.7) | 17.6 (13.0–23.9) |
| Other/no drug | 1.5 (1.5–1.5) | 1.0 (1.0–1.1) | 26.7 (24.0–29.6) | 3.1 (2.8–3.5) | 35.3 (29.0–43.1) | 3.1 (2.4–3.9) |
| C2C | 12 (1.2–1.2) | 1.1 (1.1–1.1) | 21.3 (20.1–22.6) | 2.5 (2.3–2.8) | 26.7 (23.7–30.2) | 2.4 (2.0–2.9) |
| HBP | 0.8 (0.8–0.8) | 0.9 (0.9–1.0) | 7.4 (6.9–8.0) | 2.0 (1.9–2.2) | 8.2 (7.0–9.6) | 2.0 (1.7–2.4) |
| DM | 0.8 (0.8–0.8) | 1.0 (1.0–1.0) | 7.2 (6.5–7.9) | 2.5 (2.2–2.8) | 7.5 (6.1–9.2) | 2.4 (1.9–3.0) |
| CRD | 1.3 (1.2–1.3) | 1.2 (1.2–1.2) | 7.7 (7.1–8.5) | 3.6 (3.2–4.0) | 6.7 (5.5–8.2) | 2.9 (2.3–2.6) |
| CVD | 1.1 (1.1–1.1) | 1.2 (1.1–1.2) | 10.6 (9.6–11.7) | 2.7 (2.4–3.0) | 13.1 (10.8–15.8) | 2.7 (2.2–3.5) |
| OB | 1.6 (1.6–1.7) | 1.2 (1.1–1.2) | 3.9 (3.2–4.7) | 2.7 (2.2–3.3) | 4.1 (2.7–6.0) | 3.1 (2.0–4.6) |
| DEM | 3.2 (3.0–3.3) | 3.4 (3.2–3.5) | 72.1 (62.8–82.8) | 9.9 (8.5–11.6) | 92.8 (71.8–120.0) | 10.3 (7.8–13.8) |
| Several CD | 1.4 (1.4–1.5) | 1.2 (1.1–1.2) | 43.7 (41.1–46.3) | 2.9 (2.7–3.2) | 56.9 (50.3–64.4) | 2.7 (2.2–3.3) |
| C3NC | Reference | Reference | Reference | Reference | Reference | Reference |

Abbreviations: C1R, rheumatic disease cohort; C2C, cohort with comorbidities; C3NC, cohort without comorbidities; CI, confidence interval; CRD, chronic respiratory disease; CVD, cardiovascular diseases; DEM, dementia; DM, diabetes mellitus; HBP, high blood pressure; HR, hazard ratio; MTX+, treatment with methotrexate, leflunomide and/or Janus kinase inhibitors, with or without hydroxychloroquine but no other treatment; OB, obesity; PDN, treatment with prednisone at any dose with/without other drug; PsA, psoriatic arthritis; p-y, person-year; RA, rheumatoid arthritis; RTX+, treatment with rituximab, mycophenolate mofetil, belimumab and/or cyclophosphamide with/without another drug; SLE, systemic lupus erythematosus; SpA, spondyloarthritis; VAS, vasculitis

Note 1: Models included mutually exclusive categories compared cohort C1R and C2C (model 1), rheumatic diseases and chronic diseases (model 2) and rheumatic treatments (model 3) to the cohort C3NC (reference group for all models). They were adjusted for age, sex, region, number of comorbidities, vaccination status, public health insurance status and social and material deprivation indexes

### Supplementary figure 1. Infection, hospitalization and death crude rates by cohort, disease and age group during the period A

**Infection rates per 100000 person-years**

| Age | C1R | RA | SPA | SLE | VAS | PsA | C2C | CPD | CVD | OB | DEM | C3NC |
|---|---|---|---|---|---|---|---|---|---|---|---|---|
| 18-49 | 2650 | 2320 | 2786 | 2619 | 3444 | 3555 | 3202 | 2826 | 3135 | 3749 | 2856 | 2582 |
| 50-64 | 1976 | 2106 | 1952 | 1934 | 2065 | 2189 | 2207 | 2149 | 2048 | 2642 | 3919 | 1737 |
| 65-74 | 1496 | 1713 | 1444 | 1554 | 860 | 1729 | 1225 | 1426 | 1512 | 1952 | 3939 | 697 |
| 75-84 | 2239 | 2597 | 1787 | 2170 | 2026 | 1880 | 1772 | 2463 | 2234 | 2825 | 6560 | 498 |
| 85+ | 5428 | 5335 | 6626 | 8869 | 5405 | 5212 | 4550 | 5553 | 5079 | 6612 | 10220 | 1086 |

**Hospitalization rates per 100000 person-years**

| Age | C1R | RA | SPA | SLE | VAS | PsA | C2C | CPD | CVD | OB | DEM | C3NC |
|---|---|---|---|---|---|---|---|---|---|---|---|---|
| 18-49 | 127 | 325 | 83 | 266 | * | * | 137 | 115 | 182 | 251 | * | 10 |
| 50-64 | 341 | 591 | 227 | 330 | * | 305 | 215 | 302 | 300 | 547 | 892 | 24 |
| 65-74 | 453 | 596 | 373 | 518 | 397 | 412 | 242 | 406 | 396 | 679 | 1153 | 17 |
| 75-84 | 725 | 947 | * | * | 392 | * | 513 | 884 | 758 | 1138 | 2060 | 24 |
| 85+ | 1728 | 1729 | * | 4434 | 1315 | * | 1243 | 1931 | 1512 | 2503 | 2889 | 69 |

**Death rates per 100000 person-years**

| Age | C1R | RA | SPA | SLE | VAS | PsA | C2C | CPD | CVD | OB | DEM | C3NC |
|---|---|---|---|---|---|---|---|---|---|---|---|---|
| 18-49 | * | * | * | * | * | * | 4 | * | 16 | 10 | * | * |
| 50-64 | 50 | * | * | * | * | * | 16 | 33 | 51 | 46 | * | 2 |
| 65-74 | 151 | 179 | * | * | * | * | 63 | 127 | 124 | 239 | 394 | * |
| 75-84 | 418 | 490 | * | * | * | * | 238 | 475 | 380 | 605 | 947 | * |
| 85+ | 1193 | 1214 | * | * | 1315 | * | 923 | 1387 | 1143 | 1936 | 2093 | 110 |

Abbreviations: C1R, rheumatic disease cohort; C2C, cohort with comorbidities; C3NC, cohort without comorbidities; CRD, chronic respiratory disease; CVD, cardiovascular diseases; DEM, dementia; OB, obesity; PsA, psoriatic arthritis; RA, rheumatoid arthritis; SpA, spondyloarthritis; SLE, systemic lupus erythematosus; VAS, vasculitis

* Number of events lower than 5

Note 1: Rheumatic diseases and chronic diseases are dichotomic (yes/no) not mutually exclusive categories

Note 2: Colors are presented only for illustrative purposes, defined by the median and the 25 and 75 percentiles of all age group rates of C1R, C2C and C3NC. For infection rates: <1360; 1360-1975; 1976-2615; >2615/100,000 person-years. For hospitalization rates: <47; 47-214; 215-482; >482/100,000 person-years. For death rates: <3; 3-49; 50-194; >194/100,000 person-years

### Supplementary figure 2. Infection, hospitalization and death crude rates by cohort, disease and age group during the period B

**Infection rates per 100000 person-years**

| Age | C1R | RA | SPA | SLE | VAS | PsA | C2C | CPD | CVD | OB | DEM | C3NC |
|---|---|---|---|---|---|---|---|---|---|---|---|---|
| 18-49 | 3721 | 3595 | 3613 | 4079 | 3523 | 4129 | 4082 | 3879 | 4005 | 4569 | 2935 | 3819 |
| 50-64 | 2384 | 2453 | 2310 | 2799 | 2016 | 2790 | 2600 | 2472 | 2536 | 3141 | 3148 | 2126 |
| 65-74 | 1702 | 1981 | 1252 | 1371 | 1555 | 2318 | 1425 | 1667 | 1665 | 2085 | 3293 | 897 |
| 75-84 | 2194 | 2489 | 2166 | 1974 | 2008 | 1572 | 1618 | 2138 | 2032 | 2548 | 4202 | 706 |
| 85+ | 3799 | 4400 | 2420 | 4733 | 2645 | 4020 | 3102 | 3848 | 3635 | 4828 | 6534 | 816 |

**Hospitalization rates per 100000 person-years**

| Age | C1R | RA | SPA | SLE | VAS | PsA | C2C | CPD | CVD | OB | DEM | C3NC |
|---|---|---|---|---|---|---|---|---|---|---|---|---|
| 18-49 | 187 | 404 | 72 | 350 | * | * | 220 | 180 | 308 | 455 | * | 18 |
| 50-64 | 321 | 518 | 197 | 206 | 543 | 354 | 261 | 330 | 389 | 737 | 611 | 28 |
| 65-74 | 471 | 660 | 242 | 457 | * | 724 | 253 | 418 | 418 | 711 | 861 | 15 |
| 75-84 | 794 | 926 | 825 | * | 473 | * | 398 | 699 | 620 | 990 | 1220 | 17 |
| 85+ | 1147 | 1558 | * | * | * | * | 805 | 1251 | 1049 | 1550 | 1633 | * |

**Death rates per 100000 person-years**

| Age | C1R | RA | SPA | SLE | VAS | PsA | C2C | CPD | CVD | OB | DEM | C3NC |
|---|---|---|---|---|---|---|---|---|---|---|---|---|
| 18-49 | * | * | * | * | * | * | 5 | * | 18 | 10 | * | 1 |
| 50-64 | 57 | * | * | * | * | * | 22 | 32 | 60 | 65 | 153 | 1 |
| 65-74 | 151 | 225 | * | * | * | * | 55 | 116 | 118 | 162 | 243 | 3 |
| 75-84 | 359 | 455 | * | * | * | * | 149 | 264 | 257 | 354 | 543 | * |
| 85+ | 892 | 1051 | * | * | 882 | * | 496 | 905 | 661 | 940 | 1031 | * |

Abbreviations: C1R, rheumatic disease cohort; C2C, cohort with comorbidities; C3NC, cohort without comorbidities; CRD, chronic respiratory disease; CVD, cardiovascular diseases; DEM, dementia; OB, obesity; PsA, psoriatic arthritis; RA, rheumatoid arthritis; SpA, spondyloarthritis; SLE, systemic lupus erythematosus; VAS, vasculitis

* Number of events lower than 5

Note 1: Rheumatic diseases and chronic diseases are dichotomic (yes/no) not mutually exclusive categories

Note 2: Colors are presented only for illustrative purposes, defined by the median and the 25 and 75 percentiles of all age group rates of C1R, C2C and C3NC. For infection rates: <1251; 1251-2193; 2194-3410; >3410/100,000 person-years. For hospitalization rates: <23; 23-252; 253-433; >433/100,000 person-years. For death rates: <6; 6-23; 24-149; >149/100,000 person-years

### Supplementary figure 3. Infection, hospitalization and death crude rates by cohort, disease and age group during the period C

**Infection rates per 100000 person-years**

| Age | C1R | RA | SPA | SLE | VAS | PsA | C2C | CPD | CVD | OB | DEM | C3NC |
|---|---|---|---|---|---|---|---|---|---|---|---|---|
| 18-49 | 13151 | 14418 | 12646 | 13027 | 11705 | 15052 | 11006 | 11577 | 12263 | 13101 | 12674 | 7875 |
| 50-64 | 7761 | 9379 | 6951 | 8141 | 6698 | 7158 | 5711 | 7094 | 6807 | 7796 | 15461 | 4454 |
| 65-74 | 8385 | 10253 | 7185 | 6427 | 8945 | 6266 | 4709 | 6681 | 6803 | 7996 | 23306 | 2313 |
| 75-84 | 13968 | 16458 | 11307 | 10095 | 15823 | 9472 | 8520 | 12119 | 11432 | 13342 | 31792 | 3557 |
| 85+ | 26944 | 29907 | 23725 | 22328 | 26150 | 23122 | 19138 | 24631 | 23352 | 27059 | 49054 | 8692 |

**Hospitalization rates per 100000 person-years**

| Age | C1R | RA | SPA | SLE | VAS | PsA | C2C | CPD | CVD | OB | DEM | C3NC |
|---|---|---|---|---|---|---|---|---|---|---|---|---|
| 18-49 | 348 | 348 | 188 | 321 | 1638 | 457 | 146 | 163 | 314 | 228 | | 14 |
| 50-64 | 897 | 897 | 342 | 1005 | 1725 | 449 | 262 | 553 | 573 | 686 | 1681 | 29 |
| 65-74 | 1851 | 1851 | 1255 | 1322 | 2134 | 2163 | 584 | 1297 | 1114 | 1486 | 4150 | 84 |
| 75-84 | 3682 | 3682 | 3280 | 1798 | 3370 | 3391 | 1524 | 2956 | 2424 | 3329 | 5863 | 292 |
| 85+ | 7049 | 7049 | 6069 | 7104 | 4495 | 7471 | 3783 | 6384 | 5164 | 6479 | 8863 | 1140 |

**Death rates per 100000 person-years**

| Age | C1R | RA | SPA | SLE | VAS | PsA | C2C | CPD | CVD | OB | DEM | C3NC |
|---|---|---|---|---|---|---|---|---|---|---|---|---|
| 18-49 | * | * | * | * | * | * | 9 | 9 | 24 | 13 | * | 1 |
| 50-64 | 98 | 98 | * | * | * | * | 36 | 73 | 87 | 69 | 276 | 3 |
| 65-74 | 382 | 382 | 188 | 300 | * | * | 104 | 233 | 220 | 289 | 720 | 14 |
| 75-84 | 1019 | 1019 | 1122 | 830 | 793 | 1130 | 336 | 704 | 597 | 699 | 1496 | 63 |
| 85+ | 2257 | 2257 | * | * | 1948 | * | 1349 | 2274 | 1920 | 2625 | 3663 | 352 |

Abbreviations: C1R, rheumatic disease cohort; C2C, cohort with comorbidities; C3NC, cohort without comorbidities; CRD, chronic respiratory disease; CVD, cardiovascular diseases; DEM, dementia; OB, obesity; PsA, psoriatic arthritis; RA, rheumatoid arthritis; SpA, spondyloarthritis; SLE, systemic lupus erythematosus; VAS, vasculitis

* Number of events lower than 5

Note 1: Rheumatic diseases and chronic diseases are dichotomic (yes/no) not mutually exclusive categories

Note 2: Colors are presented only for illustrative purposes, defined by the median and the 25 and 75 percentiles of all age group rates of C1R, C2C and C3NC. For infection rates: <5210; 5210-8384; 8385-12078; >12078/100,000 person-years. For hospitalization rates: <204; 204-583; 584-1686; >1686/100,000 person-years. For death rates: <11; 11-97; 98-366; >367/100,000 person-years
